# Advancing Rheumatology Practice: Systematic Review of Natural Language Processing Applications

**DOI:** 10.1101/2024.03.07.24303959

**Authors:** Mahmud Omar, Benjamin S. Glicksberg, Hagar Reuveni, Girish N. Nadkarni, Eyal Klang

## Abstract

**Background:** With the advent of large language models (LLM), such as ChatGPT, natural language processing (NLP) is revolutionizing healthcare. We systematically reviewed NLP’s role in rheumatology and assessed its impact on diagnostics, disease monitoring, and treatment strategies.

**Methods:** Following PRISMA guidelines, we conducted a systematic search to identify original research articles exploring NLP applications in rheumatology. This search was performed in PubMed, Embase, Web of Science, and Scopus until January 2024.

**Results:** Our search produced 17 studies that showcased diverse applications of NLP in rheumatology, addressing disease diagnosis, data handling, and monitoring.

Notably, GPT-4 demonstrated strong performance in diagnosing and managing rheumatic diseases. Performance metrics indicated high accuracy and reliability in various tasks. However, challenges like data dependency and limited generalizability were noted.

**Conclusion:** NLP, and especially LLM, show promise in advancing rheumatology practice, enhancing diagnostic precision, data handling, and patient care. Future research should address current limitations, focusing on data integrity and model generalizability.

## Introduction

The integration of artificial intelligence (AI) in medicine is revolutionizing healthcare (1–3). Central to this AI revolution in healthcare are Natural Language Processing (NLP) methodologies and the use of generative Large Language Models (LLM) (4–6), marked by the introduction of ChatGPT at the end of 2022. These advanced technologies have demonstrated remarkable capability in interpreting and analyzing clinical data in a human-like manner (6,7).

This technological evolution holds particular promise for rheumatology—a field grappling with a diverse range of disorders characterized by significant variability in organs involved, symptoms, treatment responses, and prognosis, which complicates patient management (8–10). NLP, known for its capability to process unstructured clinical data, is gaining recognition in rheumatology as a valuable tool, enhancing both patient care and research methodologies (11–14).

Our study aims to systematically review NLP and LLM contributions to the field of Rheumatology. This effort seeks not only to inform healthcare professionals about the benefits of NLP but also to pave the way for future research.

## Methods

### Search Strategy

This systematic review was registered with the International Prospective Register of Systematic Reviews - PROSPERO (Registration code CRD42024509490). We adhered to the Preferred Reporting Items for Systematic Reviews and Meta-Analyses (PRISMA) guidelines (15,16).

A systematic search was conducted across PubMed, Embase, Web of Science, and Scopus, up until January 2024. We complemented the search via reference screening for any additional papers. We aimed to identify original research articles that investigated the application of NLP and LLM in the diagnosis or prediction of rheumatological diseases.

The search utilized a combination of keywords including “Natural Language Processing”, “NLP”, “Large Language Models”, “LLMs”, “Artificial Intelligence Models”, “AI Models”, “Rheumatology”, “Rheumatologic Diseases”, “Rheumatoid Arthritis”, “Systemic Lupus Erythematosus”, “Sjogren’s Syndrome”, “Scleroderma”, “Polymyositis”, “Dermatomyositis”, “Ankylosing Spondylitis”, “Psoriatic Arthritis”, “Gout”, “Osteoarthritis”, “Data Analysis”, “Predictive Modeling”, “Pattern Recognition”, “Text Mining”, “Electronic Health Records”, “EHR Analysis”, “Diagnosis”, and “Prediction”.

Specific search strings for each database are detailed in the Supplementary Materials, tailored to PubMed, Embase, Scopus, and Web of Science. The search strings employed in each database varied slightly to optimize the retrieval of relevant articles, encompassing a broad spectrum of studies that intersect the fields of natural language processing, and various rheumatologic diseases.

### Study Selection

We included original research articles that focused on the application of NLP and LLM in diagnosing, classifying, or predicting rheumatic diseases. Studies were selected if they provided data for assessing the performance metrics of AI models, such as area under the curve, accuracy, sensitivity, and specificity. We excluded review papers, case reports, conference abstracts, editorials, preprints, and studies not conducted in English. We also excluded studies employing AI techniques unrelated to NLP.

### Data Extraction

Two independent reviewers extracted relevant information using a standardized form. Data points included year of publication, study design, sample size, specific conversational NLP techniques used, dataset details for model training and validation, performance metrics, and key findings. Discrepancies between reviewers were resolved through discussion, and a third reviewer was consulted when necessary.

### Risk of Bias

To evaluate the quality and robustness of the methodologies in the included studies, the Quality Assessment Tool for Observational Cohort and Cross-Sectional Studies tool was used (17).

## Results

### Search Results and Study Selection

The process of study selection and the screening methodology are detailed in the PRISMA flow chart (**Figure 1**). Our search yielded a total of 691 articles, with a breakdown of 106 from PubMed, 213 from Embase, 246 from Scopus, and 126 from Web of Science. Following the removal of 402 duplicates, 289 articles remained for title and abstract screening. This process led to the exclusion of 226 articles, narrowing the field to 63 full-text articles for thorough evaluation.

**Figure 1:**
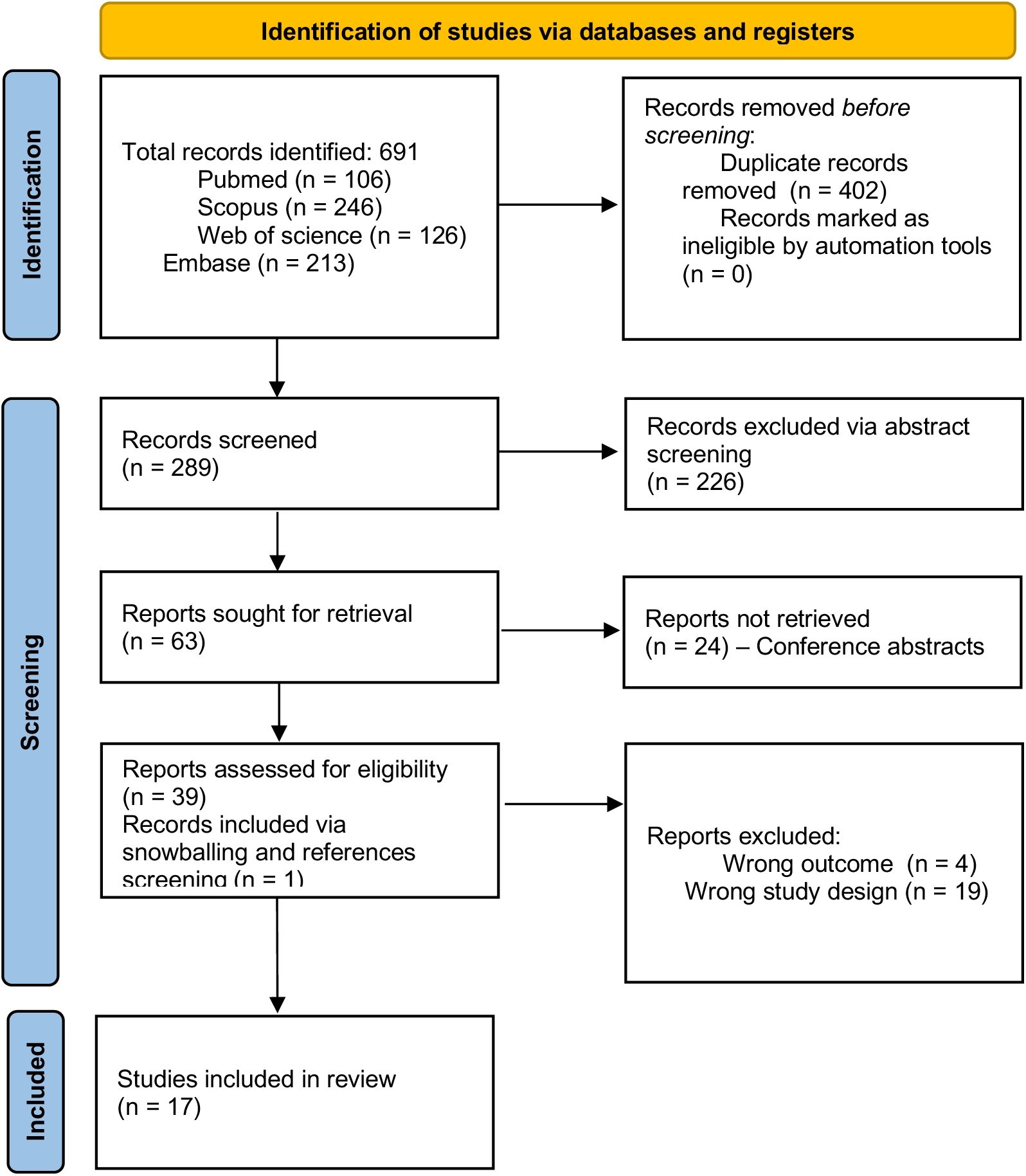
PRISMA flowchart.

Ultimately, 16 studies were chosen for inclusion based on their relevance and adherence to our criteria. One additional study was identified and included through reference screening (11–14,18–30). Therefore, the review culminated in a total of 17 studies. The included studies were published between 2010 and 2024 (**Figure 1**).

### Quality assessment

We assessed the quality of the included 17 studies using the Quality Assessment Tool for Observational Cohort and Cross-Sectional Studies (17), excluding criteria not applicable to our study designs. Most studies clearly stated their research questions, defined study populations, and employed consistent selection criteria. However, a common gap was the lack of justification for sample sizes and power descriptions.

While exposure measures were typically clear and valid, not all studies measured exposure before the outcome, affecting their quality ratings. Additionally, the blinding of outcome assessors and adjustment for key confounding variables were frequently overlooked. Despite these issues, the overall quality of the studies varied from ‘Fair’ to ‘Good’, with only one study rated as ‘Poor’ **(Table 1).**

**Table 1:** Quality Assessment Tool for Observational Cohort and Cross-Sectional Studies.

| Study (Ref) | Q1 | Q2 | Q3 | Q4 | Q5 | Q6 | Q7 | Q8 | Q9 | Q10 | Q11 | overall |
| --- | --- | --- | --- | --- | --- | --- | --- | --- | --- | --- | --- | --- |
| Benavent et al.(18) | yes | yes | yes | No | yes | yes | NA | yes | yes | No | No | Fair |
| Osborne et al.(19) | yes | yes | yes | No | yes | yes | NA | yes | yes | No | NA | Good |
| Zhao et al. (20) | yes | yes | yes | No | yes | NA | NA | yes | yes | NA | No | Fair |
| Luedders et al.(21) | yes | yes | yes | No | yes | yes | NA | yes | yes | No | NA | Good |
| Liao et al. (24) | yes | yes | yes | No | yes | yes | NA | yes | yes | No | NA | Good |
| Humbert-Droz et al. (23) | yes | yes | yes | No | no | NA | yes | yes | Yes | No | yes | Good |
| Yoshida et al.(24) | yes | yes | yes | No | yes | NA | NA | yes | yes | No | No | Fair |
| England et al. (25) | yes | yes | yes | No | yes | yes | NA | yes | yes | No | yes | Good |
| Love et al. (26) | yes | yes | yes | No | yes | yes | yes | yes | yes | NA | yes | Good |
| Zheng et al. (14) | yes | yes | yes | No | yes | NA | NA | yes | yes | No | No | Fair |
| Krusche et al.(13) | yes | yes | yes | No | no | NA | NA | yes | yes | Yes | No | Poor |
| Gräf et al. (27) | yes | yes | yes | yes | yes | NA | NA | yes | yes | yes | no | Fair |
| Wang et al. (28) | yes | yes | yes | yes | yes | NA | NA | yes | yes | yes | yes | Good |
| Lin et al. (12) | yes | yes | yes | yes | yes | NA | NA | yes | yes | yes | no | Fair |
| Walsh et al. (29) | yes | yes | yes | yes | yes | NA | NA | yes | yes | yes | yes | Good |
| Redd et al.(30) | yes | yes | yes | yes | yes | NA | NA | yes | yes | yes | yes | Good |
| van Leeuwen et al. (11) | yes | yes | yes | No | yes | yes | NA | yes | yes | CD | yes | Good |

Abbreviations:
Q1: Research Question Clearly Stated | Q2: Study Population Clearly Specified and Defined | Q3: Subjects
Selected from Similar Populations with Uniform Criteria | Q4: Sample Size Justification and Power Description |
Q5: Exposure Measured Prior to Outcome | Q6: Sufficient Timeframe to See an Association | Q7: Examination of
Different Levels of Exposure | Q8: Clear, Valid, Reliable Exposure Measures | Q9: Outcome Measures Clearly
Defined, Valid, Reliable | Q10: Blinding of Outcome Assessors to Exposure Status | Q11: Adjustment for Key
Potential Confounding Variable.

### Summary of the included studies

The included 17 studies highlight the evolving application of NLP and language models like ChatGPT and BERT in rheumatology (**Table 2**). These studies, encompassing sample sizes ranging from a few hundred to over 34 million clinical notes, underscore the role of NLP in various rheumatologic conditions, including axial spondyloarthritis, psoriatic arthritis, rheumatoid arthritis, gout, and systemic sclerosis. The clinical tasks addressed were diverse, covering disease activity assessment, condition identification, and diagnostic accuracy enhancements **(Table 3).** Importantly, the studies primarily focused on the use of NLP and language models, often supplementing them with tabular machine learning techniques to improve outcomes **(Figure 2)**. Performance metrics, such as precision, recall, F1 scores, and area under the curve, generally pointed to a high level of accuracy and reliability in the application of NLP and language models **(Tables 3-4).**

**Figure 2:**
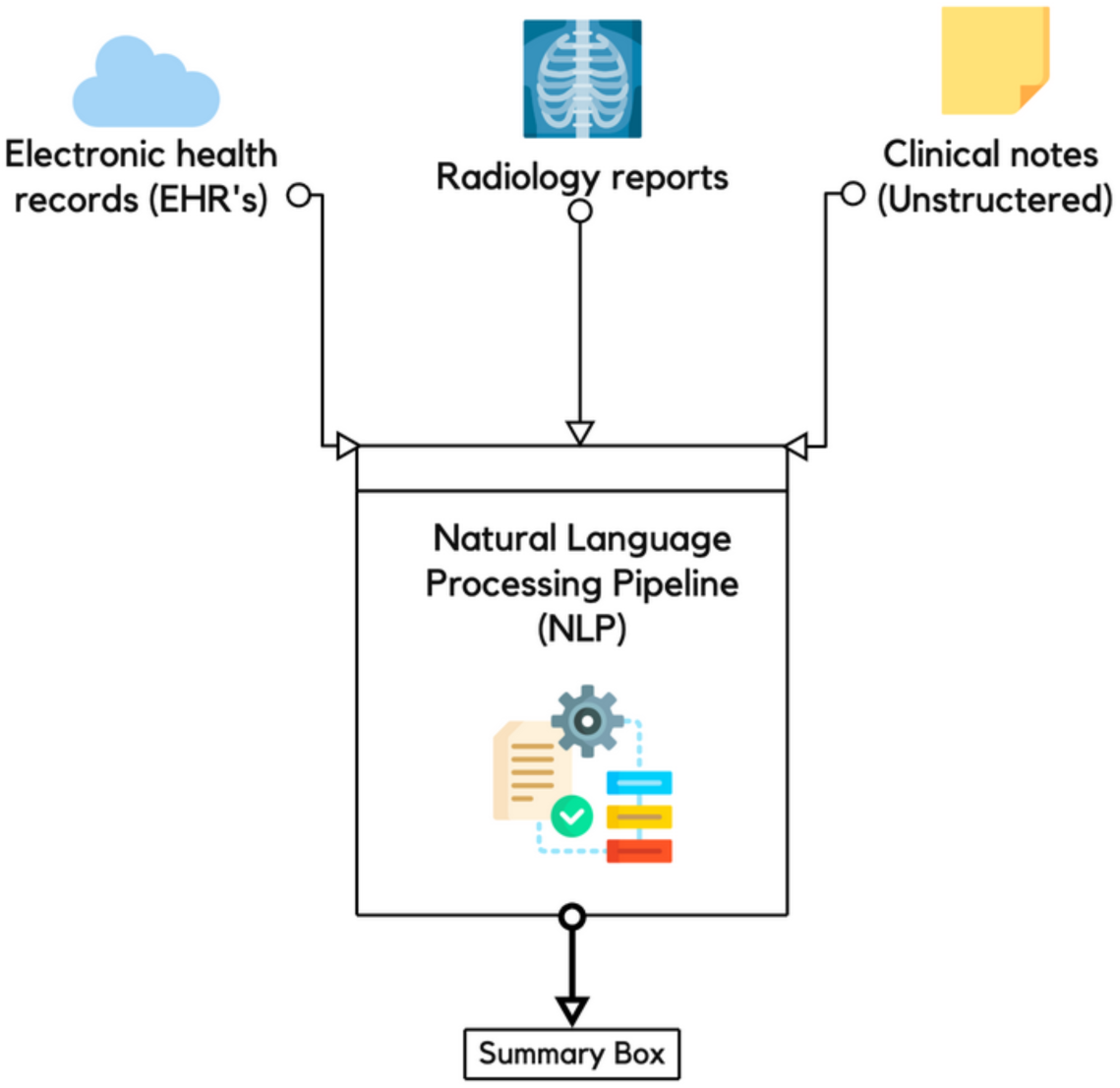
Simple Diagnostic Pathway Using NLP Analysis.

**Table 2:**
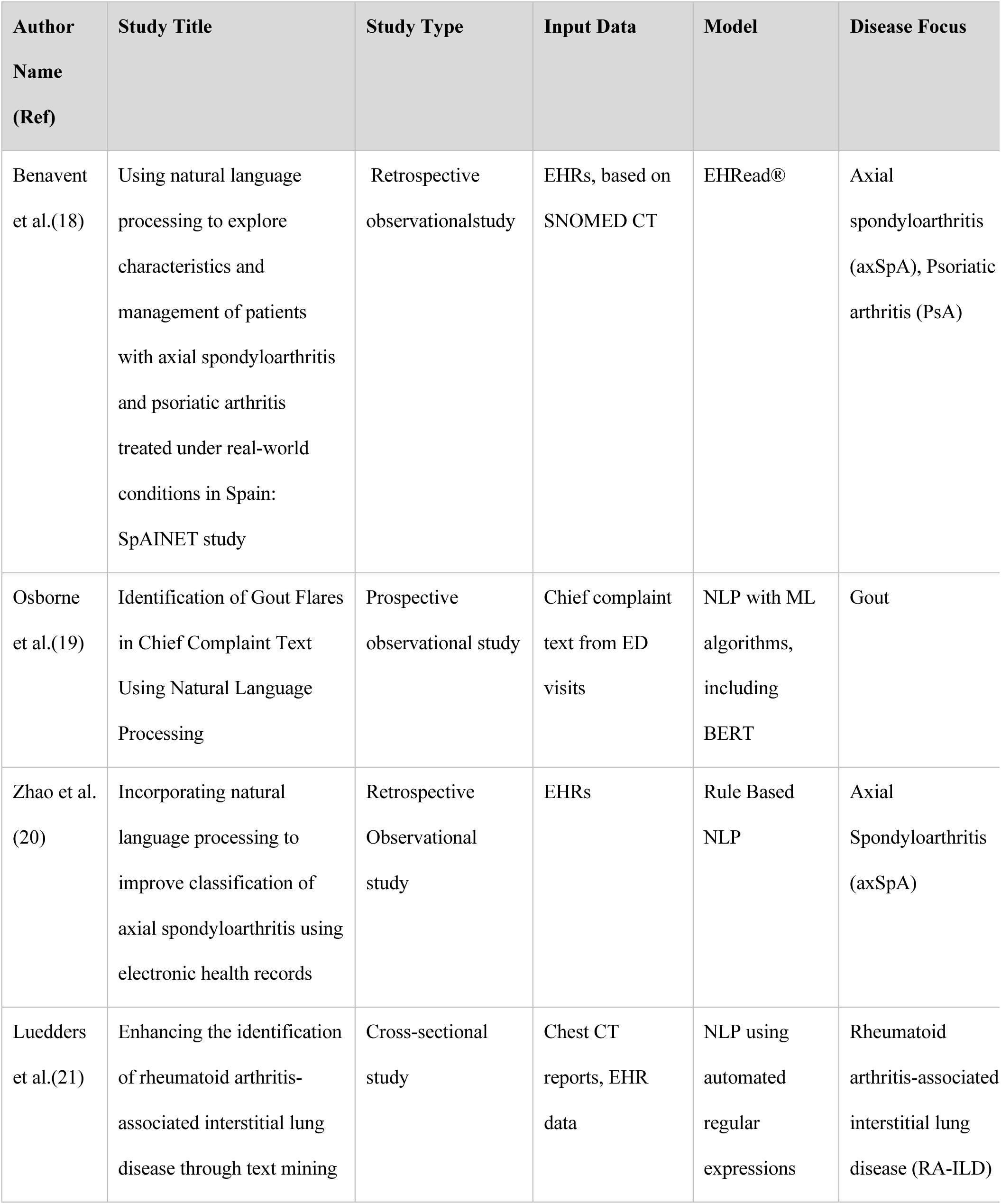
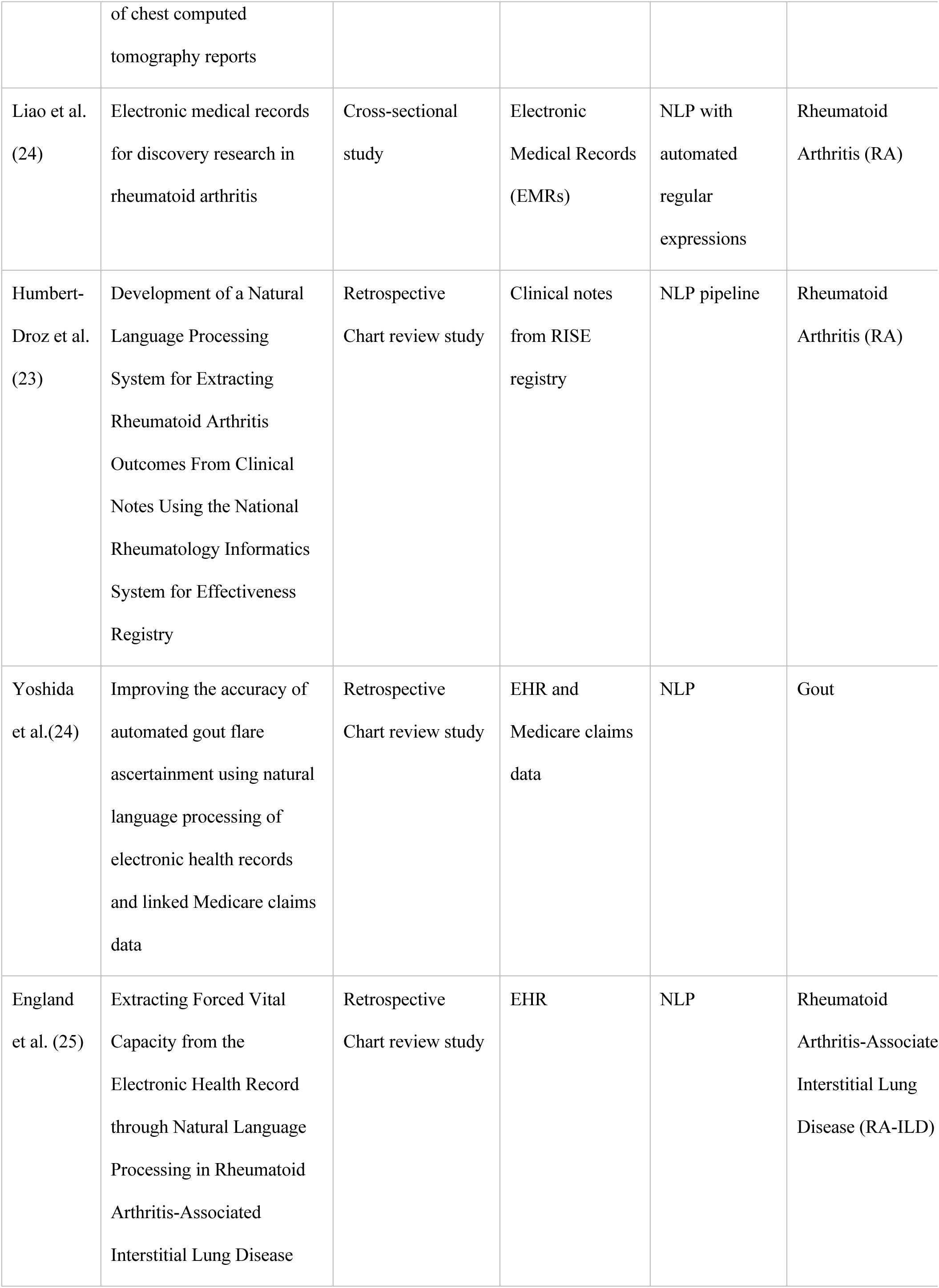
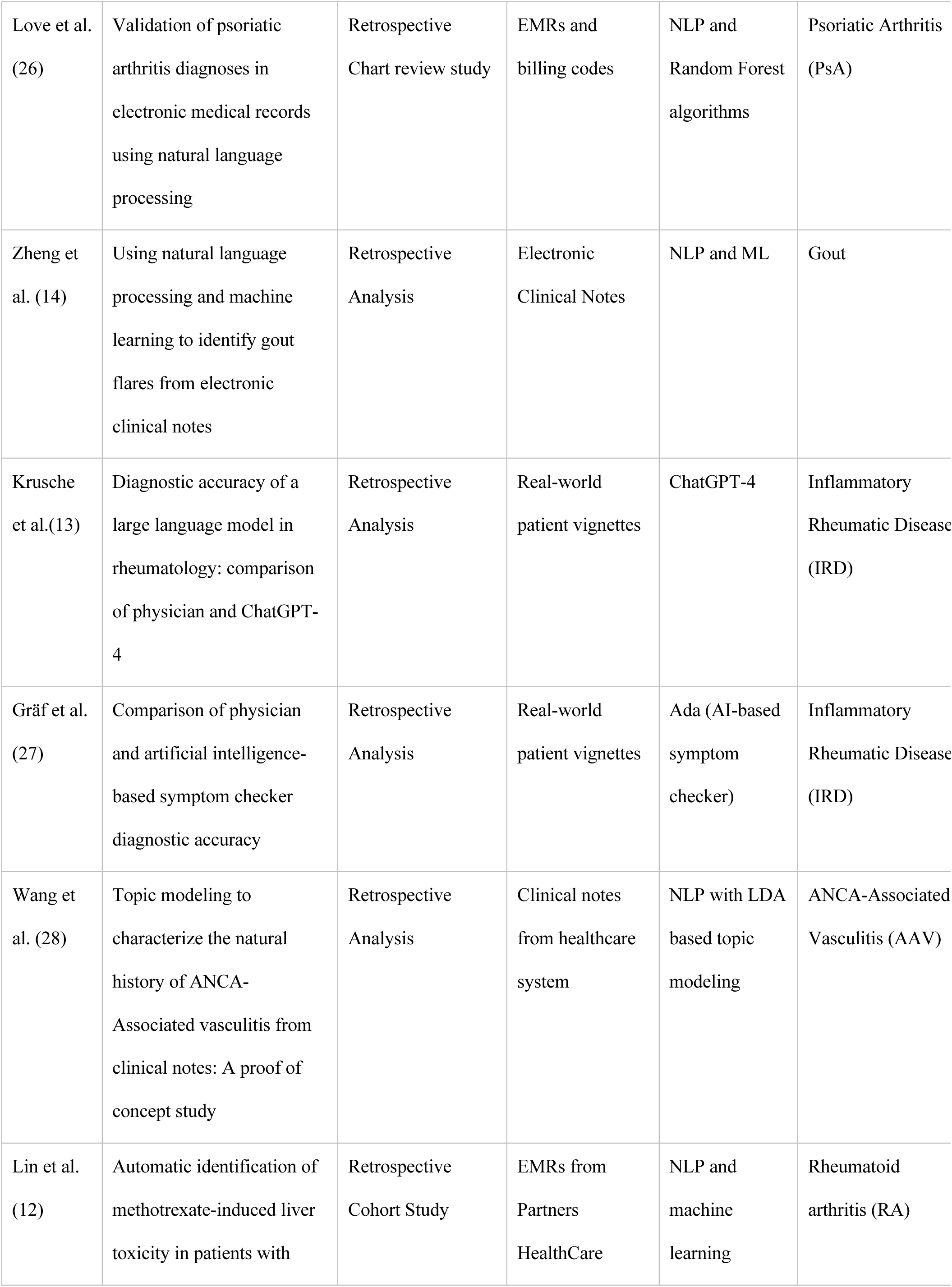
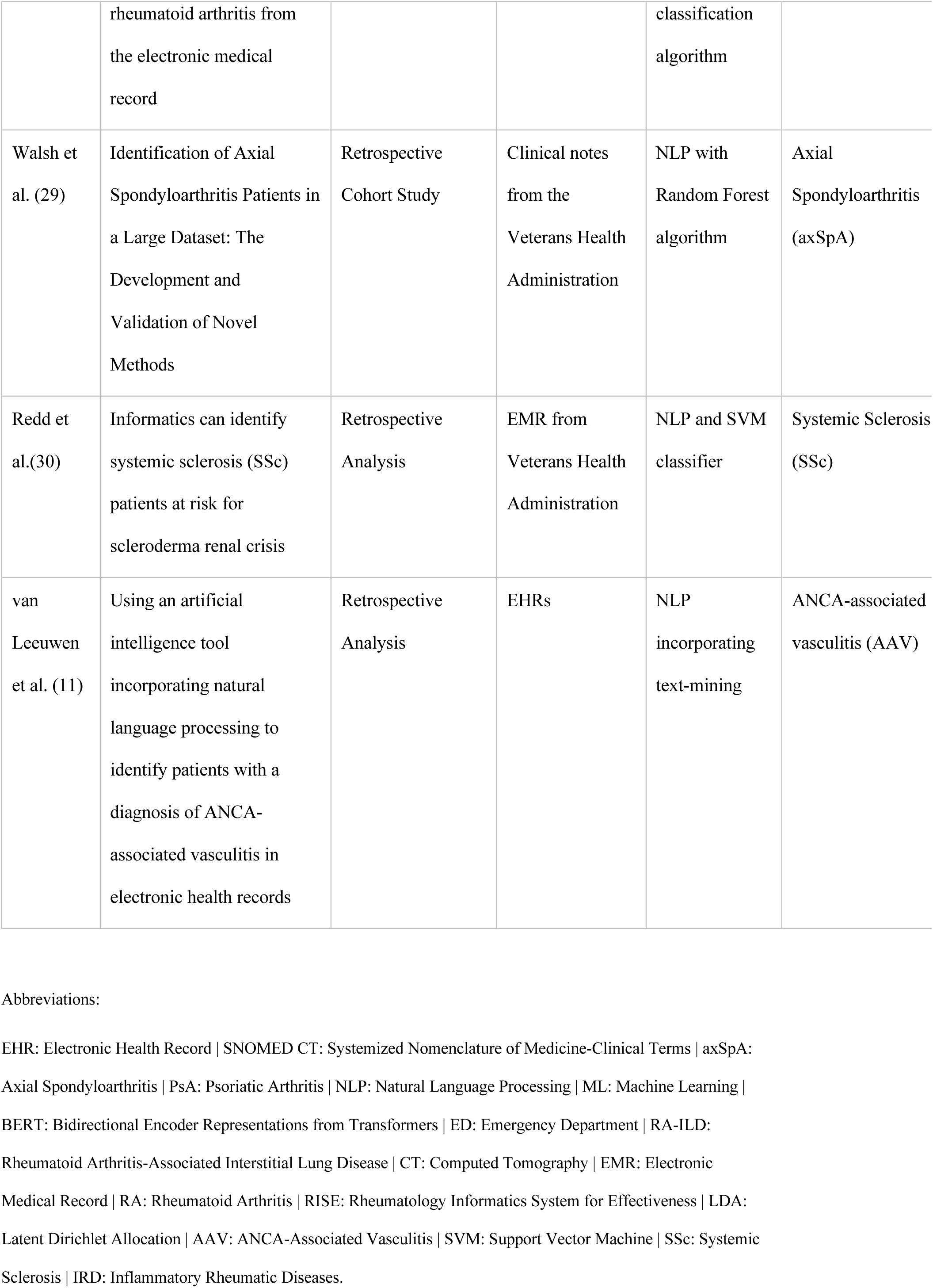
Summary of the included studies.

**Table 3:**
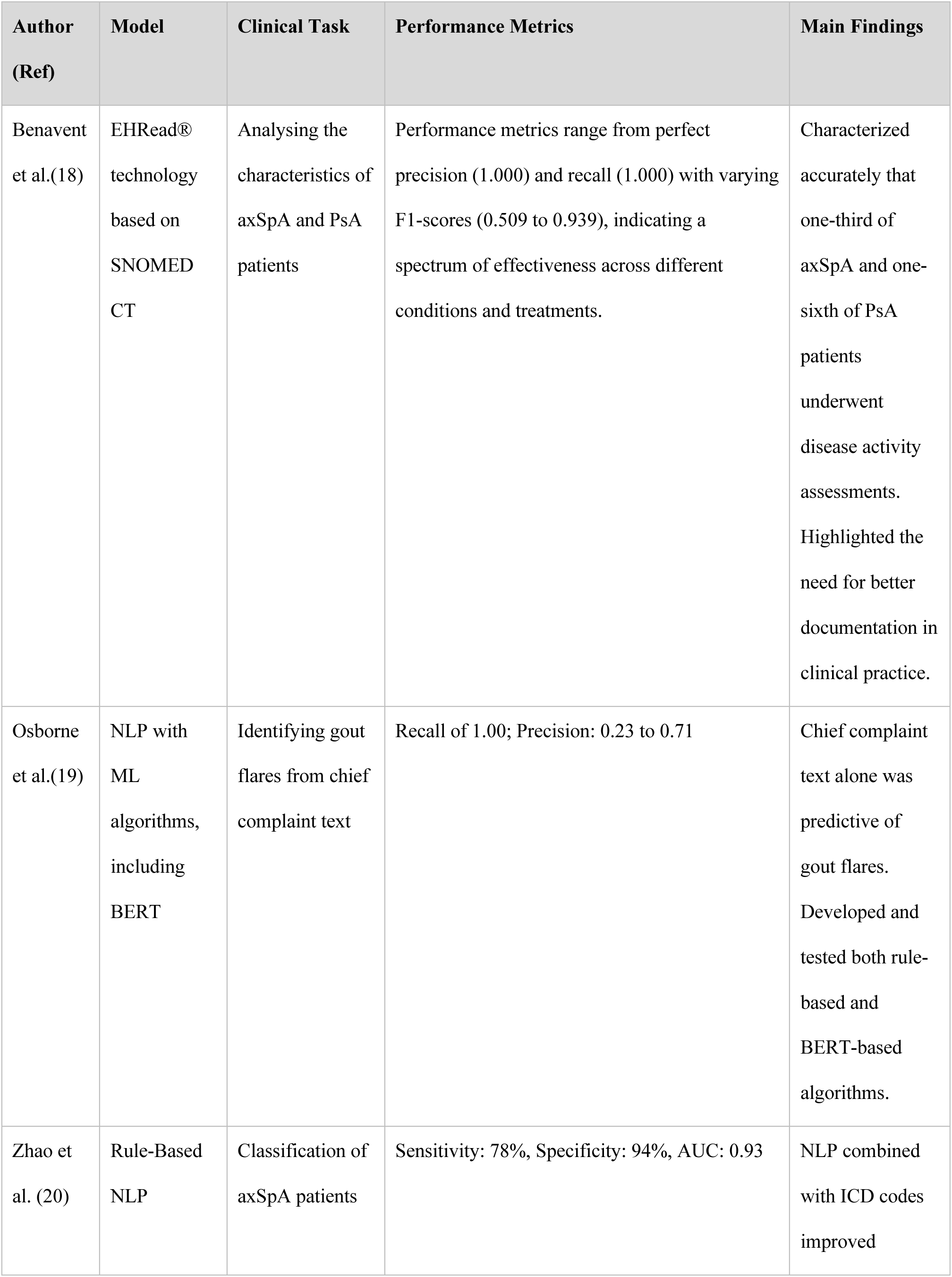
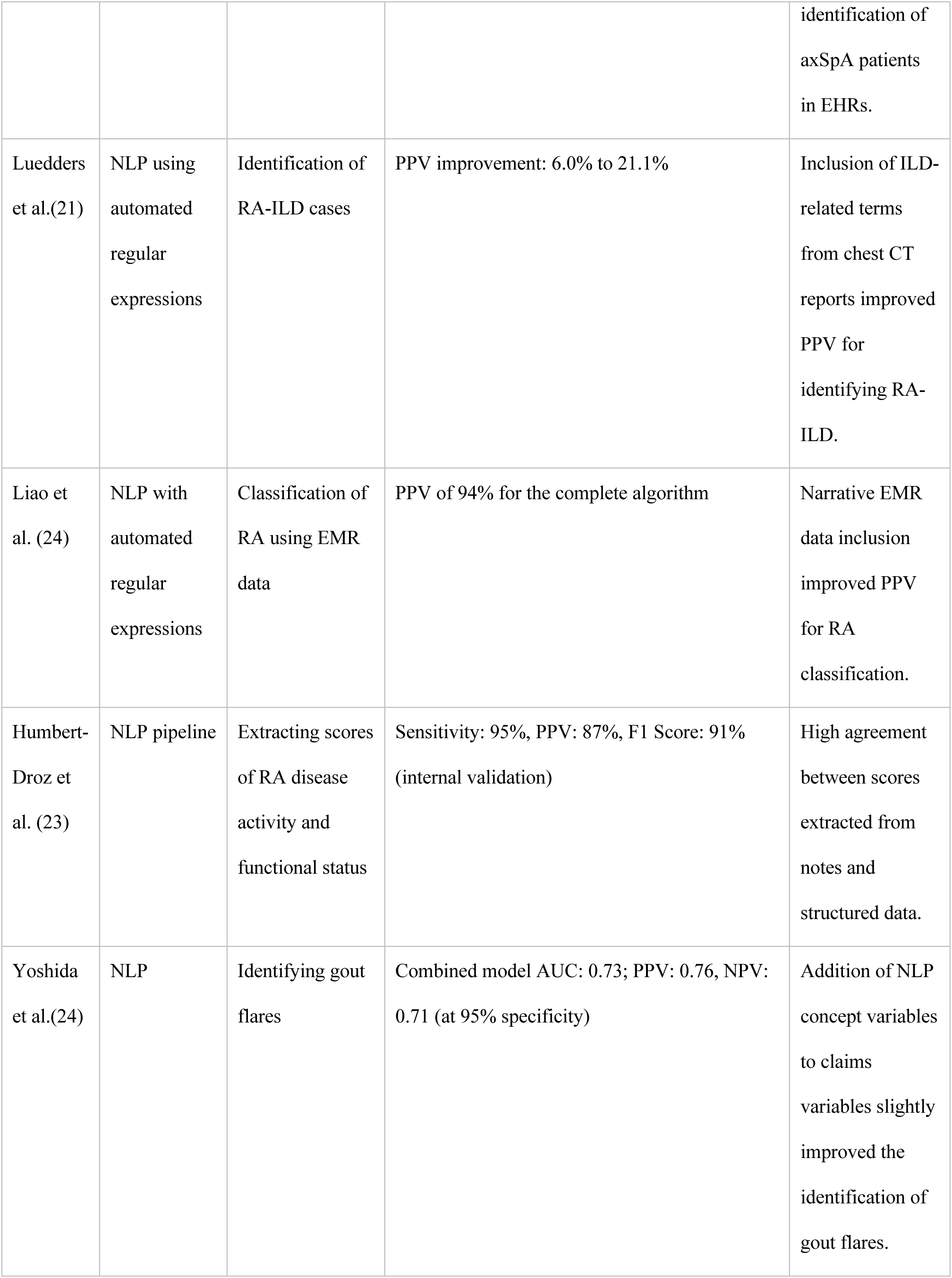
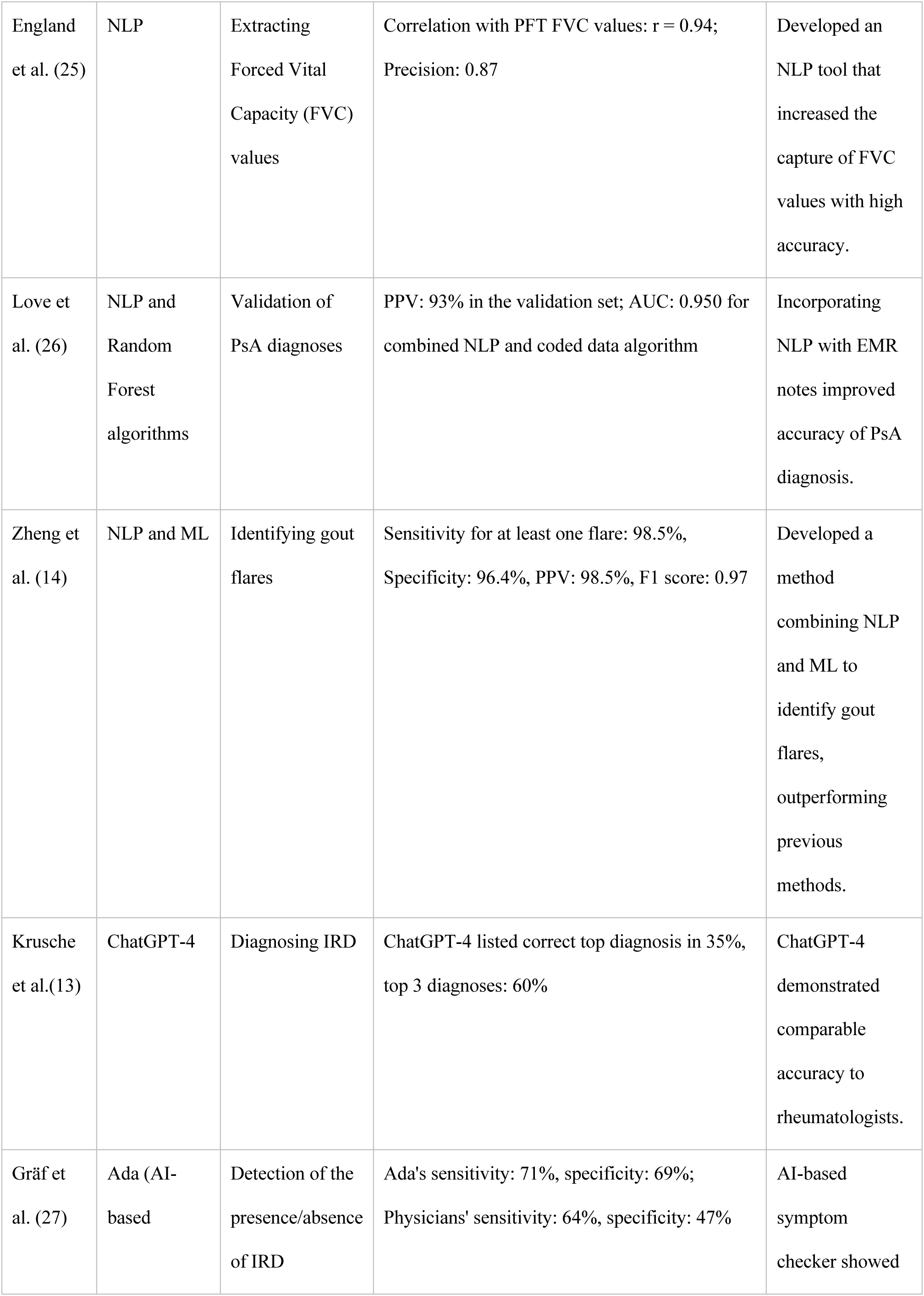
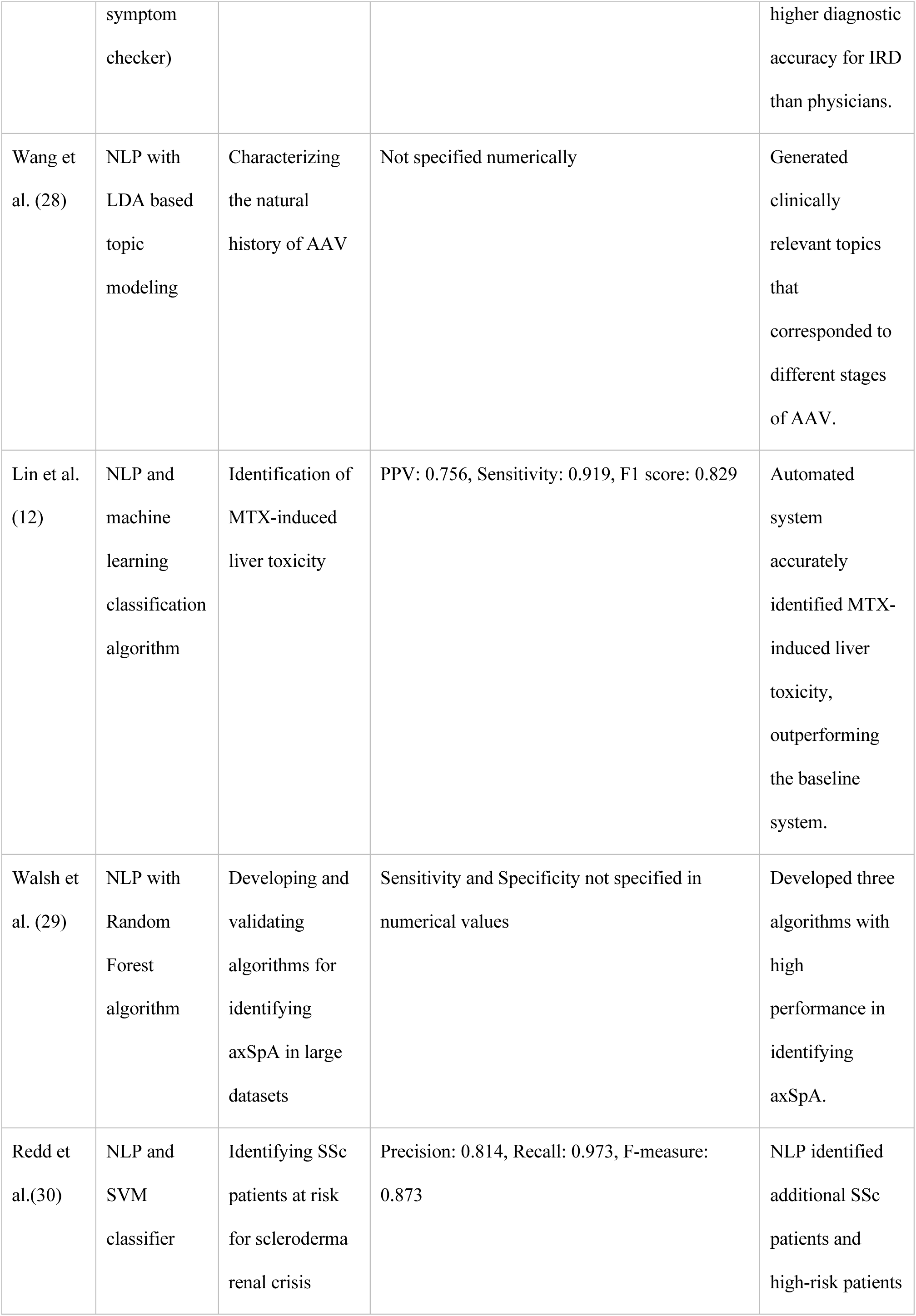
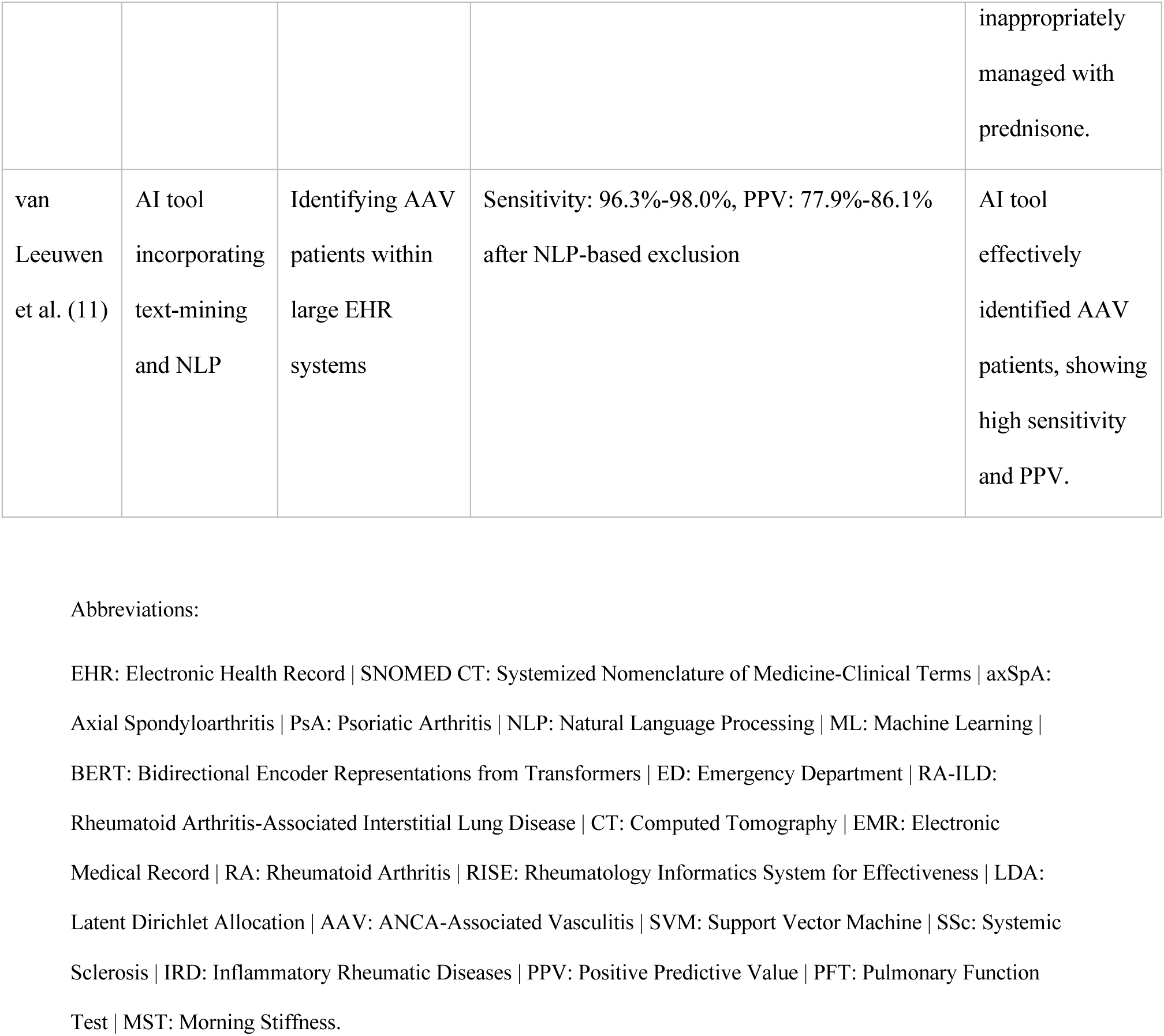
Performance Metrics and Key Findings.

**Table 4:**
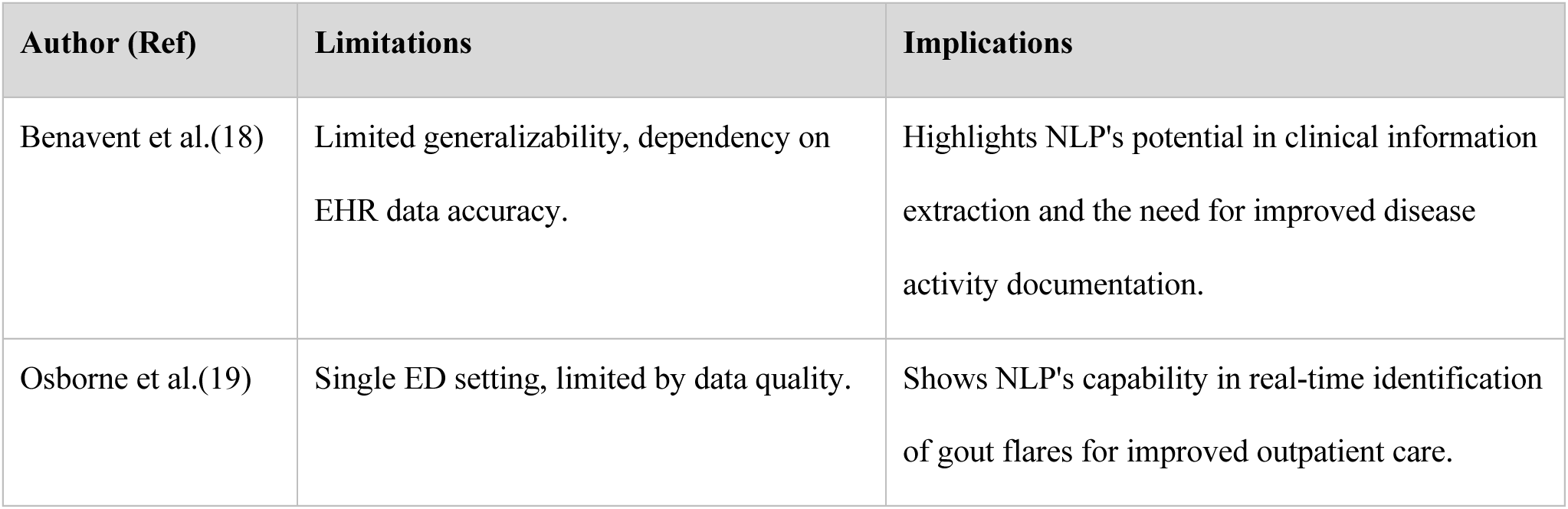
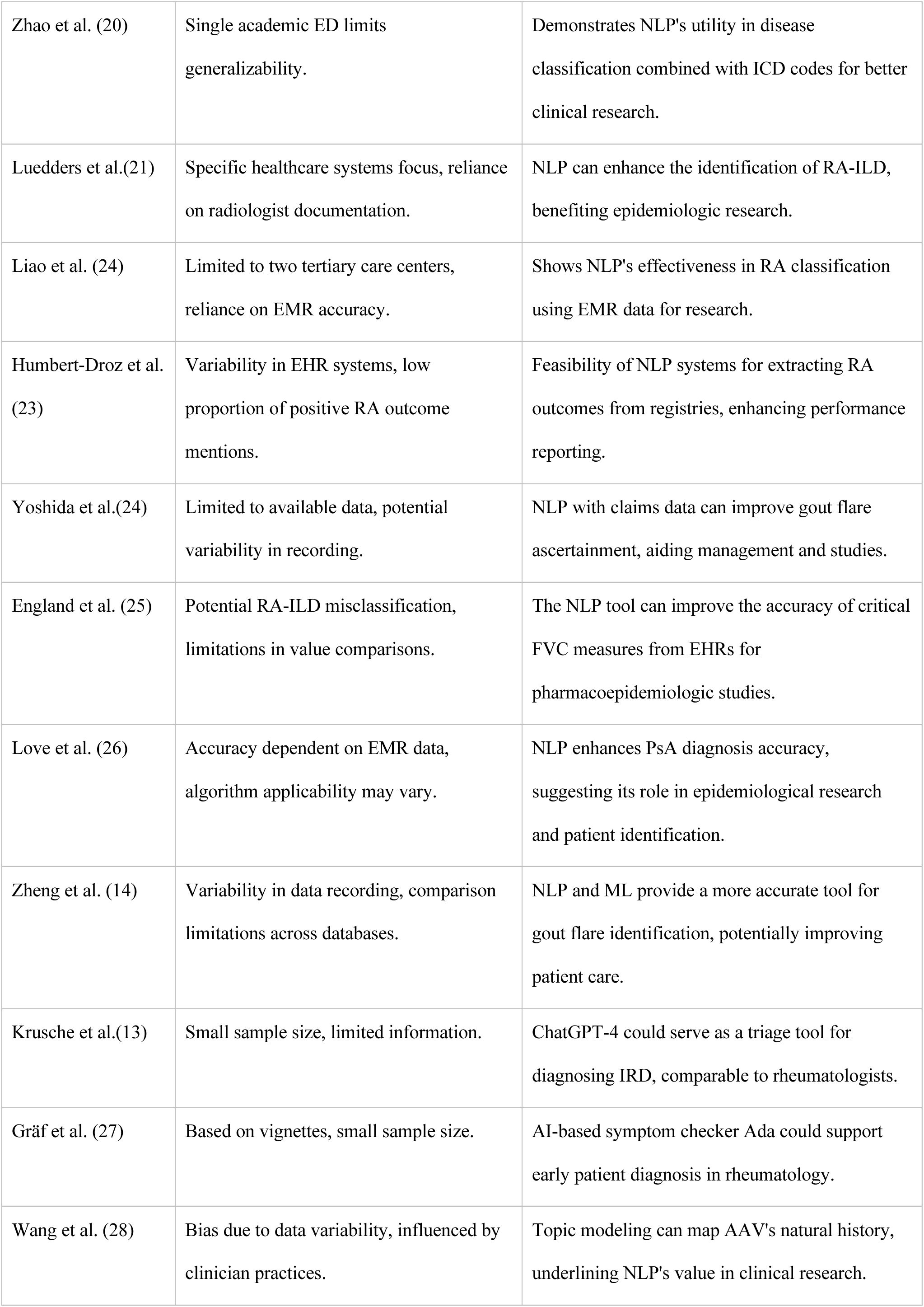
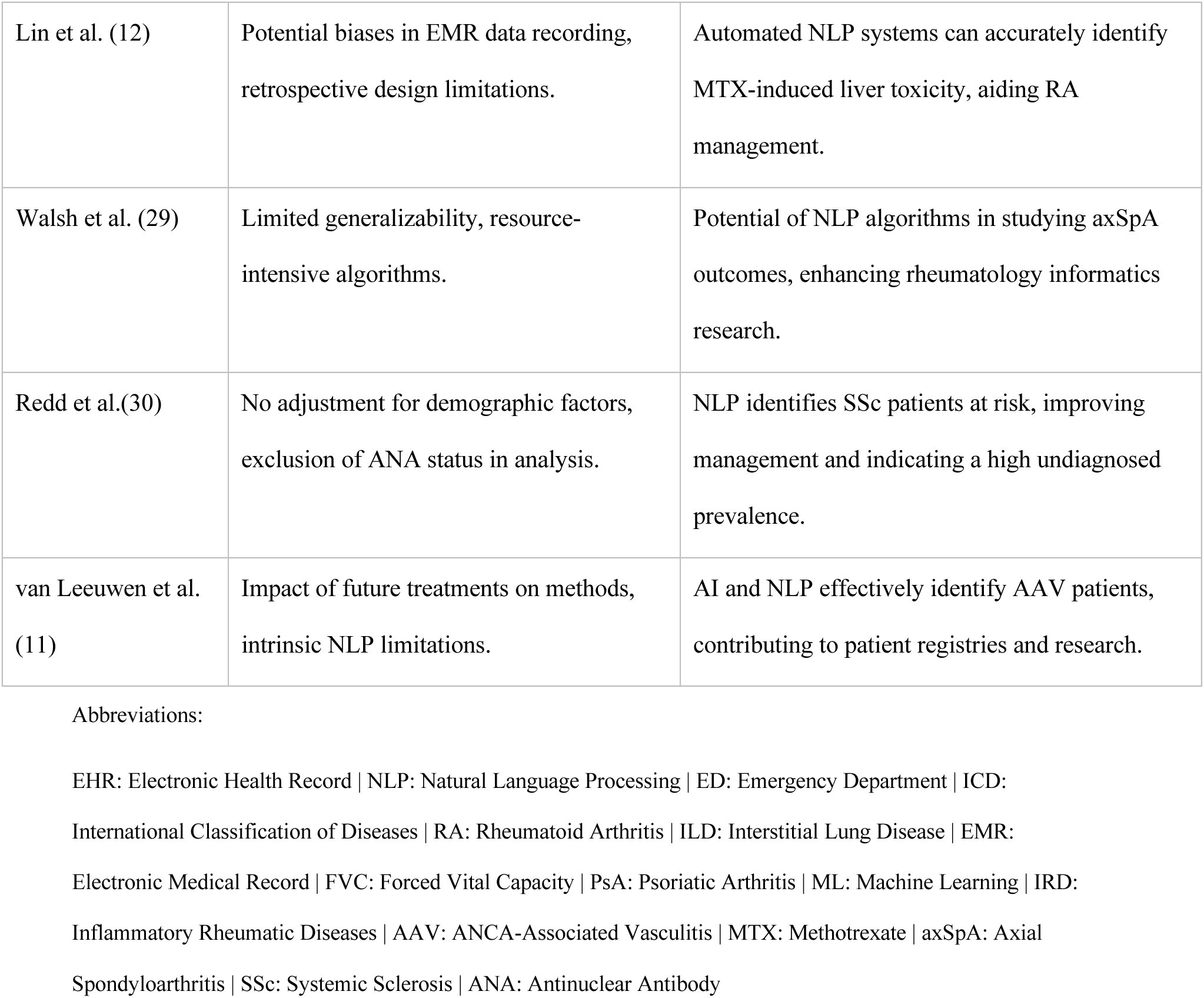
Limitations and Implications.

### NLP tasks in Rheumatology Research

#### Disease Diagnosis and Classification

Significant strides in rheumatology diagnosis and classification are evident through studies utilizing NLP. Krusche et al. (2024) employed GPT-4 in diagnosing inflammatory rheumatic diseases, achieving a correct top diagnosis in 35% of cases (13). In a similar vein, van Leeuwen et al. (2024) effectively identified ANCA-associated vasculitis using an AI tool with NLP, showing a sensitivity range of 96.3% to 98.0% (11). Additionally, Love et al. (2011) improved the accuracy of psoriatic arthritis diagnoses in EMRs using NLP, achieving a PPV of 93% (26). These studies demonstrate the precision and reliability of NLP in disease identification and classification. Zhao et al. (2020) also contributed to this area by improving the identification of axial spondyloarthritis, sacroiliitis, and HLA-B27 positive patients using EHRs, with their unsupervised algorithm achieving a sensitivity of 78% and specificity of 94% (20). Humbert-Droz et al. (2023) exemplified the use of NLP in analyzing large-scale clinical data, extracting Rheumatoid Arthritis outcomes from over 34 million notes with high sensitivity (95%) and PPV (87%) (23). Walsh et al. (2020) developed algorithms to identify Axial Spondyloarthritis with impressive accuracy. Their Spond NLP algorithm particularly stood out for its sensitivity and specificity, illustrating the potential of NLP in early disease detection and risk assessment (29).

#### Disease Activity Assessment and Management

NLP has shown promise in advancing disease activity assessment and management **(Figure 3)**.

**Figure 3:**
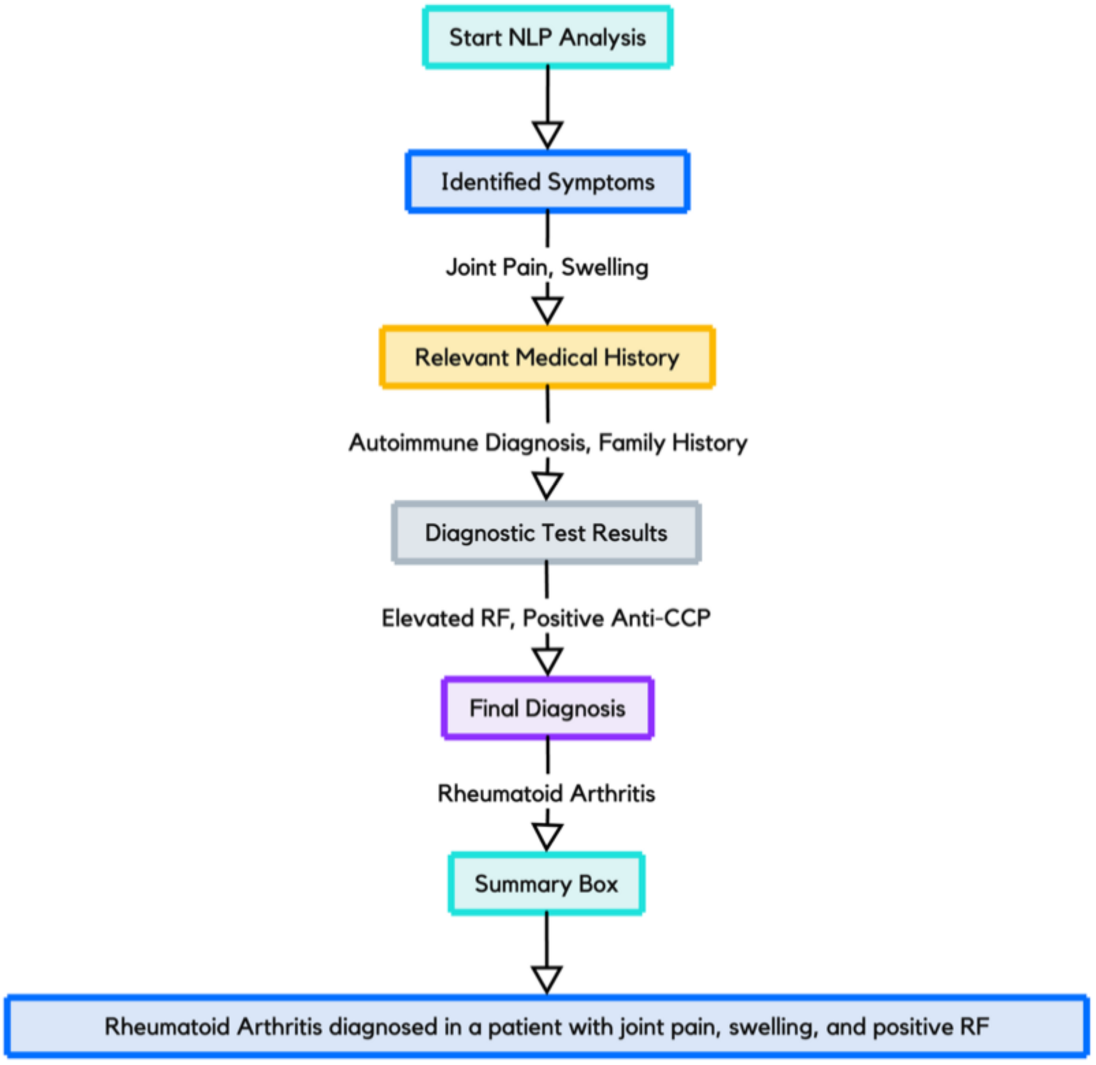
An Example of NLP-Enhanced Diagnostic Workflow for Clinical Decision Support.

The SpAINET study by Benavent et al. (2023) utilized NLP to manage axial spondyloarthritis and psoriatic arthritis, with notable precision in disease activity assessment (Etanercept precision score of 1.000) (18). England et al. (2024) developed a NLP tool for extracting Forced Vital Capacity from EHRs, demonstrating high correlation (r = 0.94) with pulmonary function test values, thus underlining the potential of NLP in enhancing patient care through precise data analysis (25).

#### Predictive Modeling and Risk Assessment

Redd et al. (2014) utilized NLP to identify systemic sclerosis patients at risk for renal crisis, underscoring the role of NLP in predictive health analytics (30). Additionally, Lin et al. (2015) focused on identifying methotrexate-induced liver toxicity in patients with rheumatoid arthritis using NLP and machine learning classification algorithms (12). Their approach achieved a positive predictive value of 0.756, highlighting NLP’s capability to anticipate treatment-related complications, an essential aspect of patient safety and personalized care.

## Discussion

Our systematic review reveals a potential for integration of NLP and LLMs in rheumatology. These models can improve diagnostics, disease monitoring, and treatment strategies across various rheumatic disorders.

NLP has the potential to transform rheumatology by enhancing patient care and research (9). With the rise of digital patient health records and advanced diagnostics, there’s a surge in patient data. AI methods, including NLP, machine learning, and deep learning, are pivotal in harnessing this data for predicting outcomes and guiding clinical decisions (1,8). In rheumatology, AI models have significantly improved the diagnosis of diseases like rheumatoid arthritis using various models (9,10,31). These models aid in screening, disease identification, patient phenotyping in EHRs, assessing treatment responses, and monitoring disease progression (31,32).

Additionally, AI contributes to risk assessment for comorbidities, drug discovery, and advancing basic science research, making it a powerful tool in modern rheumatology practice (8,9,31–34). Research has shown NLP has practical applications in rheumatology. Studies like Osborne et al.’s (2021) on identifying gout flares using NLP in emergency settings, and Krusche et al.’s (2024) work on employing GPT-4 for diagnosing inflammatory rheumatic diseases, showcase NLP’s potential in enhancing diagnostic accuracy (13,19).

The application of LLMs, such as GPT-4, appears to hold significant potential. In Krusche et al.’s study, GPT-4 not only demonstrated remarkable accuracy in diagnosing various rheumatic diseases, but also surpassed even expert rheumatologists in precision (achieving 60% accuracy in the top 3 diagnoses compared to 55% by rheumatologists) (13). Despite the study’s limited sample size, these findings indicate promise for LLMs in enhancing triage and diagnostic processes in routine clinical practice.

Similarly, Zhao et al.’s (2020) improvement in identifying axial spondyloarthritis patients and Humbert-Droz et al.’s (2023) extraction of rheumatoid arthritis outcomes from clinical notes demonstrate NLP’s effectiveness in disease classification and management (20,23).

Despite the benefits, the application of NLP in rheumatology comes with limitations. Challenges include data quality dependency, small sample sizes, and the need for wider generalizability in studies (9). Additionally, the complexity of rheumatic diseases necessitates sophisticated and adaptable models, which can accurately reflect diseases’ heterogeneity (8,9).

In conclusion, NLP, and especially LLM, show promise in advancing rheumatology practice, enhancing diagnostic precision, data handling, and patient care. Future research should address current limitations, focusing on data integrity and model generalizability.

## Data Availability

All data produced in the present study are available upon reasonable request to the authors

## Acknowledgment

none

## Supplementary materials

The Boolean strings used for each database:

**Pubmed**: (“Natural Language Processing” OR “NLP” OR “Large Language Models” OR “LLMs” OR “Artificial Intelligence Models” OR “AI Models”) AND (“Rheumatology” OR “Rheumatologic Diseases” OR “Rheumatoid Arthritis” OR “Systemic Lupus Erythematosus” OR “Sjogren’s Syndrome” OR “Scleroderma” OR “Polymyositis” OR “Dermatomyositis” OR “Ankylosing Spondylitis” OR “Psoriatic Arthritis” OR “Gout” OR “Osteoarthritis”) AND (“Data Analysis” OR “Predictive Modeling” OR “Pattern Recognition” OR “Text Mining” OR “Electronic Health Records” OR “EHR Analysis” OR “Diagnosis” OR “Prediction”)

**Embase**: (’natural language processing’ OR ‘nlp’ OR ‘large language models’ OR ‘llms’ OR ‘artificial intelligence models’ OR ‘ai models’) AND (’rheumatology’ OR ‘rheumatologic diseases’ OR ‘rheumatoid arthritis’ OR ‘systemic lupus erythematosus’ OR ‘sjogren’s syndrome’ OR ‘scleroderma’ OR ‘polymyositis’ OR ‘dermatomyositis’ OR ‘ankylosing spondylitis’ OR ‘psoriatic arthritis’ OR ‘gout’ OR ‘osteoarthritis’) AND (’data analysis’ OR ‘predictive modeling’ OR ‘pattern recognition’ OR ‘text mining’ OR ‘electronic health records’ OR ‘ehr analysis’ OR ‘diagnosis’ OR ‘prediction’)

**Scopus**: (TITLE-ABS-KEY (“Natural Language Processing” OR “NLP” OR “Large Language Models” OR “LLMs” OR “AI” OR “Artificial Intelligence”) AND TITLE- ABS-KEY (“Rheumatology” OR “Rheumatoid Arthritis” OR “Systemic Lupus Erythematosus” OR “Sjogren&apos;s Syndrome” OR “Scleroderma” OR “Polymyositis” OR “Dermatomyositis” OR “Ankylosing Spondylitis” OR “Psoriatic Arthritis”) AND TITLE-ABS-KEY (“Machine Learning” OR “Predictive Models” OR “Text Mining” OR “EHR Analysis”))

**Web of Science**: (TS=(“Natural Language Processing” OR “NLP” OR “Large Language Models” OR “LLMs” OR “AI” OR “Artificial Intelligence”) AND TS=(“Rheumatology” OR “Rheumatoid Arthritis” OR “Systemic Lupus Erythematosus” OR “Sjogren’s Syndrome” OR “Scleroderma” OR “Polymyositis” OR “Dermatomyositis” OR “Ankylosing Spondylitis” OR “Psoriatic Arthritis”) AND TS=(“Machine Learning” OR “Predictive Models” OR “Text Mining” OR “EHR Analysis”))

## Notes

### Competing Interest Statement

The authors have declared no competing interest.

### Funding Statement

This study did not receive any funding

### Author Declarations

The study used (or will use) ONLY openly available human data that were originally located at:

## References

1. Rajpurkar P, Chen E, Banerjee O, Topol EJ. AI in health and medicine. Nat Med. 2022 Jan 20;28(1):31–8.

2. Davenport T, Kalakota R. The potential for artificial intelligence in healthcare. Future Healthc J. 2019 Jun;6(2):94–8.

3. Beam AL, Drazen JM, Kohane IS, Leong TY, Manrai AK, Rubin EJ. Artificial Intelligence in Medicine. New England Journal of Medicine. 2023 Mar 30;388(13):1220–1.

4. Wilhelm TI, Roos J, Kaczmarczyk R. Large Language Models for Therapy Recommendations Across 3 Clinical Specialties: Comparative Study. J Med Internet Res. 2023 Oct 30;25:e49324.

5. Nadkarni PM, Ohno-Machado L, Chapman WW. Natural language processing: an introduction. Journal of the American Medical Informatics Association. 2011 Sep 1;18(5):544–51.

6. Chary M, Parikh S, Manini A, Boyer E, Radeous M. A Review of Natural Language Processing in Medical Education. Western Journal of Emergency Medicine. 2018 Dec 12;20(1):78–86.

7. Feller DJ, Zucker J, Yin MT, Gordon P, Elhadad N. Using Clinical Notes and Natural Language Processing for Automated HIV Risk Assessment. JAIDS Journal of Acquired Immune Deficiency Syndromes. 2018 Feb 1;77(2):160–6.

8. McMaster C, Bird A, Liew DFL, Buchanan RR, Owen CE, Chapman WW, et al. Artificial Intelligence and Deep Learning for Rheumatologists. Arthritis & Rheumatology. 2022 Dec 26;74(12):1893–905.

9. Chinnadurai S, Mahadevan S, Navaneethakrishnan B, Mamadapur M. Decoding Applications of Artificial Intelligence in Rheumatology. Cureus. 2023 Sep 28;

10. Momtazmanesh S, Nowroozi A, Rezaei N. Artificial Intelligence in Rheumatoid Arthritis: Current Status and Future Perspectives: A State-of-the-Art Review. Rheumatol Ther. 2022 Oct 18;9(5):1249–304.

11. van Leeuwen JR, Penne EL, Rabelink T, Knevel R, Teng YKO. Using an artificial intelligence tool incorporating natural language processing to identify patients with a diagnosis of ANCA-associated vasculitis in electronic health records. Comput Biol Med. 2024 Jan;168:107757.

12. Lin C, Karlson EW, Dligach D, Ramirez MP, Miller TA, Mo H, et al. Automatic identification of methotrexate-induced liver toxicity in patients with rheumatoid arthritis from the electronic medical record. Journal of the American Medical Informatics Association. 2015 Apr 1;22(e1):e151–61.

13. Krusche M, Callhoff J, Knitza J, Ruffer N. Diagnostic accuracy of a large language model in rheumatology: comparison of physician and ChatGPT-4. Rheumatol Int. 2023 Sep 24;44(2):303–6.

14. Zheng C, Rashid N, Wu Y, Koblick R, Lin AT, Levy GD, et al. Using Natural Language Processing and Machine Learning to Identify Gout Flares From Electronic Clinical Notes. Arthritis Care Res (Hoboken). 2014 Nov 24;66(11):1740–8.

15. Page MJ, McKenzie JE, Bossuyt PM, Boutron I, Hoffmann TC, Mulrow CD, et al. The PRISMA 2020 statement: an updated guideline for reporting systematic reviews. BMJ. 2021 Mar 29;n71.

16. Schiavo JH. PROSPERO: An International Register of Systematic Review Protocols. Med Ref Serv Q. 2019 Apr 3;38(2):171–80.

17. Ma LL, Wang YY, Yang ZH, Huang D, Weng H, Zeng XT. Methodological quality (risk of bias) assessment tools for primary and secondary medical studies: what are they and which is better? Mil Med Res. 2020 Dec 29;7(1):7.

18. Benavent D, Muñoz-Fernández S, De la Morena I, Fernández-Nebro A, Marín-Corral J, Castillo Rosa E, et al. Using natural language processing to explore characteristics and management of patients with axial spondyloarthritis and psoriatic arthritis treated under real-world conditions in Spain: SpAINET study. Ther Adv Musculoskelet Dis. 2023 Jan 24;15.

19. Osborne JD, Booth JS, O’Leary T, Mudano A, Rosas G, Foster PJ, et al. Identification of Gout Flares in Chief Complaint Text Using Natural Language Processing. AMIA Annu Symp Proc. 2020;2020:973–82.

20. Zhao SS, Hong C, Cai T, Xu C, Huang J, Ermann J, et al. Incorporating natural language processing to improve classification of axial spondyloarthritis using electronic health records. Rheumatology. 2020 May 1;59(5):1059–65.

21. Luedders BA, Cope BJ, Hershberger D, DeVries M, Campbell WS, Campbell J, et al. Enhancing the identification of rheumatoid arthritis-associated interstitial lung disease through text mining of chest computerized tomography reports. Semin Arthritis Rheum. 2023 Jun;60:152204.

22. Liao KP, Cai T, Gainer V, Goryachev S, Zeng-treitler Q, Raychaudhuri S, et al. Electronic medical records for discovery research in rheumatoid arthritis. Arthritis Care Res (Hoboken). 2010 Aug 3;62(8):1120–7.

23. Humbert-Droz M, Izadi Z, Schmajuk G, Gianfrancesco M, Baker MC, Yazdany J, et al. Development of a Natural Language Processing System for Extracting Rheumatoid Arthritis Outcomes From Clinical Notes Using the National Rheumatology Informatics System for Effectiveness Registry. Arthritis Care Res (Hoboken). 2023 Mar 31;75(3):608–15.

24. Yoshida K, Cai T, Bessette LG, Kim E, Lee SB, Zabotka LE, et al. Improving the accuracy of automated gout flare ascertainment using natural language processing of electronic health records and linked Medicare claims data. Pharmacoepidemiol Drug Saf. 2024 Jan 31;33(1).

25. England BR, Roul P, Yang Y, Hershberger D, Sayles H, Rojas J, et al. Extracting forced vital capacity from the electronic health record through natural language processing in rheumatoid arthritis-associated interstitial lung disease. Pharmacoepidemiol Drug Saf. 2024 Jan 19;33(1).

26. Love TJ, Cai T, Karlson EW. Validation of Psoriatic Arthritis Diagnoses in Electronic Medical Records Using Natural Language Processing. Semin Arthritis Rheum. 2011 Apr;40(5):413–20.

27. Gräf M, Knitza J, Leipe J, Krusche M, Welcker M, Kuhn S, et al. Comparison of physician and artificial intelligence-based symptom checker diagnostic accuracy. Rheumatol Int. 2022 Sep 10;42(12):2167–76.

28. Wang L, Miloslavsky E, Stone JH, Choi HK, Zhou L, Wallace ZS. Topic modeling to characterize the natural history of ANCA-Associated vasculitis from clinical notes: A proof of concept study. Semin Arthritis Rheum. 2021 Feb;51(1):150–7.

29. Walsh JA, Pei S, Penmetsa G, Hansen JL, Cannon GW, Clegg DO, et al. Identification of Axial Spondyloarthritis Patients in a Large Dataset: The Development and Validation of Novel Methods. J Rheumatol. 2020 Jan;47(1):42–9.

30. Redd D, Frech TM, Murtaugh MA, Rhiannon J, Zeng QT. Informatics can identify systemic sclerosis (SSc) patients at risk for scleroderma renal crisis. Comput Biol Med. 2014 Oct;53:203–5.

31. Gilvaz VJ, Reginato AM. Artificial intelligence in rheumatoid arthritis: potential applications and future implications. Front Med (Lausanne). 2023 Nov 16;10.

32. Adams LC, Bressem KK, Ziegeler K, Vahldiek JL, Poddubnyy D. Artificial intelligence to analyze magnetic resonance imaging in rheumatology. Joint Bone Spine. 2024 May;91(3):105651.

33. Foulquier N, Redou P, Le Gal C, Rouvière B, Pers JO, Saraux A. Pathogenesis-based treatments in primary Sjogren’s syndrome using artificial intelligence and advanced machine learning techniques: a systematic literature review. Hum Vaccin Immunother. 2018 May 17;1–6.

34. Bellando-Randone S, Russo E, Venerito V, Matucci-Cerinic M, Iannone F, Tangaro S, et al. Exploring the Oral Microbiome in Rheumatic Diseases, State of Art and Future Prospective in Personalized Medicine with an AI Approach. J Pers Med. 2021 Jun 30;11(7):625.

